# Smaller Infarct Size with Ticagrelor vs. Clopidogrel in STEMI Patients: Insights from Cardiac MRI

**DOI:** 10.1101/2025.01.08.25320224

**Authors:** Francisco A Fonseca, Adriano Caixeta, Gilberto Szarf, Ibraim Pinto, Antonio Figueiredo-Neto, Carolina França, Henrique Bianco, Henrique Fonseca, Amanda Bacchin, Michelle Birtche, Igor Batista, Maria C Izar, the BATTLE-AMI investigators

## Abstract

**BACKGROUND:** Ticagrelor has many protective cardiovascular properties in addition to its potent antiplatelet action. This study aims to compare the effects of ticagrelor versus clopidogrel on infarcted mass, quantified by cardiac magnetic resonance imaging (cMRI), in patients with ST-segment elevation acute myocardial infarction (STEMI).

**METHODS AND RESULTS:** Adult patients of both sexes with STEMI under pharmaco-invasive strategy were included (n=225). Patients were thrombolized within six hours of symptom onset and underwent angiography and percutaneous coronary interventions, when needed, within the first 24 hours. Before the invasive procedures, patients were randomized to be treated with ticagrelor or clopidogrel (central computerized system). Patients were followed weekly to optimize medical therapy. After 30 days, cMRI was performed and smaller percentage of left ventricular infarcted mass was found with ticagrelor (p=0.012), despite similar angiographic characteristics at baseline (SINTAX score, Gensini score, culprit artery, TIMI flow or myocardial blush). At 30 days, left ventricular ejection fraction (LVEF) was comparable between groups, but the K-means algorithm displayed more homogeneous responses for smaller infarcted mass and better LVEF among those patients treated with ticagrelor. Standard lipid panel and most inflammatory parameters were similar at baseline and after 30 days. However, lower high-sensitivity troponin T and high-sensitivity C-reactive protein levels were found in samples collected from patients treated with ticagrelor, in the first day of STEMI.

**CONCLUSIONS:** In patients with STEMI under pharmaco-invasive strategy, therapy with ticagrelor was associated with smaller infarct size than clopidogrel.

**REGISTRATION:** URL:https//clinicaltrials.gov; Unique identifier: NCT02428374.

**Graphical abstract:** STEMI-ST-elevation myocardial infarction; OMT- Optimal Medical Treatment; cMRI-cardiac magnetic resonance imaging; hsTNT-high-sensitivity troponin T; hsCRP-high-sensitivity C-reactive protein.

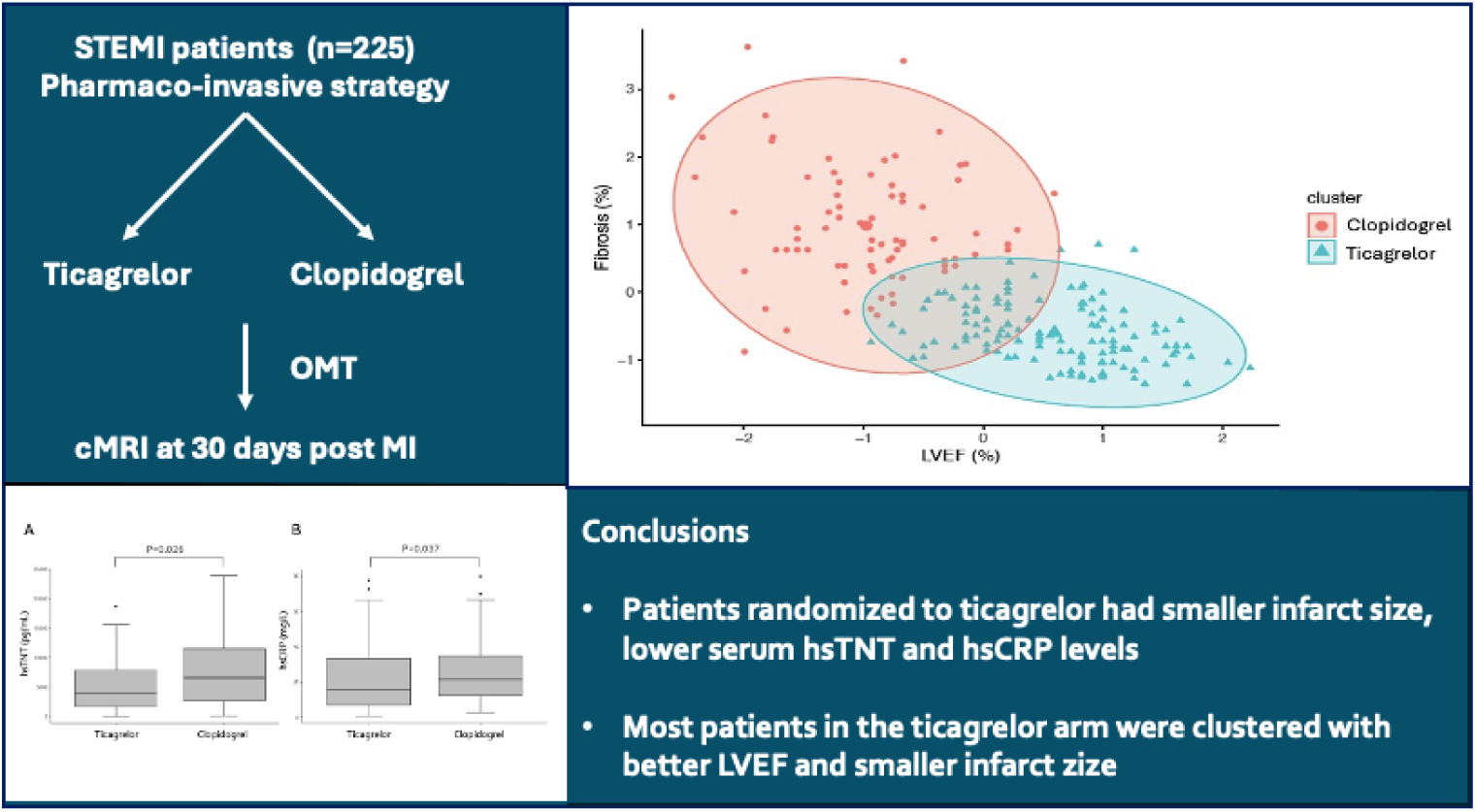

**CLINICAL PERSPECTIVE:** *What Is New?:*

- In patients with STEMI under timely pharmaco-invasive strategy, ticagrelor was associated with smaller infarct size by cMRI at day 30 compared with clopidogrel.
- This beneficial effect was related to lower serum levels of hsTNT and hsCRP at day 1.
- Randomized patients had similar pre- and post-reperfusion angiographic characteristics, and received optimal medical therapy.

*What Are the Clinical Implications?:*

- Our findings suggest greater benefit of ticagrelor, possibly related to early cardiomyocyte recovery after reperfusion in STEMI patients.
- Improvement in microcirculation and attenuation in the inflammatory response may have contributed to the findings.
- Further studies are needed to investigate the underlying mechanisms and potential clinical implications of these findings.

---

Even after more than a decade of the PLATO study^1^, the superior clinical results of ticagrelor over clopidogrel remain not fully understood. Improvements in microcirculation and enhanced antiplatelet activity have been suggested^2,3^, but the advantages of ticagrelor over clopidogrel remain controversial ^4–6^. Recently, interest in ticagrelor has been renewed due to its excellent outcomes as monotherapy in patients undergoing percutaneous coronary intervention (PCI) after an initial period of dual antiplatelet therapy.^7,8^ Additionally, in patients aged 75 years or younger, bleeding rates have been comparable to those observed with clopidogrel, even among patients receiving thrombolytic therapy^9,10^.

Currently, residual lipid^11,12^, inflammatory^13–15^, and thrombotic^16–18^ risks have been suggested as key factors contributing to the high rate of recurrent events following acute myocardial infarction^19^. Furthermore, cardiac magnetic resonance imaging (cMRI) provides an accurate assessment of infarct size and ventricular remodeling after acute myocardial infarction, offering valuable prognostic insights^20^.

In many countries, the majority of myocardial infarctions are initially managed in hospitals without 24-hour access to hemodynamic services. As a result, the pharmaco-invasive strategy becomes the most practical therapeutic option when timely PCI is not achievable^21–23^.

This prospective, randomized clinical trial aimed to compare the effects of ticagrelor and clopidogrel on infarct size and ventricular function, assessed via cMRI, in patients with ST-segment elevation myocardial infarction (STEMI) treated with a pharmaco-invasive strategy. Additionally, the study investigated potential underlying mechanisms, including inflammation, lipid profiles, and microcirculation.

## METHODS

### Study design

The study design, including its objectives and inclusion/exclusion criteria, was previously published^24^. Briefly, the BATTLE-AMI study (NCT02428374) is a prospective, randomized, open-label clinical trial with blinded cMRI and angiographic analyses. Adult STEMI patients who underwent thrombolytic therapy were included. After providing written informed consent, eligible patients were randomized in a 1:1 ratio to receive either ticagrelor or clopidogrel using a centralized computerized system prior to coronary angiography.

### Patient Recruitment and Treatment

Consecutive STEMI patients of both sexes, aging 18-75 years, were treated by tenecteplase (Metalyse^®^, Boehringer Ingelheim) in the first 6 hours of symptom onset and received 300 mg of clopidogrel and 300 mg of aspirin. They were referred to our university hospital in the first 24 hours, and randomly assigned 1:1 to either ticagrelor (Brilinta^®^, Astrazeneca) 90 mg b.i.d. or clopidogrel (Plavix^®^, Sanofi) 75 mg q.d.

Coronary angiography was performed in all patients in the first 24 hours and PCI, when needed, in the culprit lesion. Additional coronary interventions in non-culprit lesions were performed at the same time or electively according to the hemodynamic team decision. Patients with clinical instability, previous myocardial infarction or coronary revascularization, severe renal insufficiency, active liver disease, immune diseases, or known intolerance to the study drugs, were excluded. cMRI study was performed 30 days after myocardial infarction. Patients were included from May 2015 to March 2020. The study protocol was approved by local ethics committee (IRB 0297/2014; CAAE: 71652417.3.0000.5505) which follows the last version of Helsinki Declaration. All patients signed the informed consent prior to any study procedure. All laboratory, angiographic or MRI analyses were performed blindly.

### Laboratory analysis

Blood samples were collected in the first day, or in the morning of the next day in case of overnight hospital admission, and also after 30 days.

Routine laboratory parameters were performed in the central laboratory of the university hospital. B and T Lymphocyte subtypes were examined by flow-cytometry as previously reported^25^. Circulating cytokines were determined by enzyme-linked immunosorbent assay (ELISA)^26^.

### Quantitative Coronary Angiography

An independent angiographic core laboratory assessed quantitative coronary angiography (QCA) blinded to randomization assignment and clinical outcomes for baseline and post-PCI lesions with the use of validated quantitative methods (CMS; Medis Medical Imaging Systems, Leiden, The Netherlands).^27,28^ The path line, vessel contour, lumen, reference diameters, and lesion length were determined automatically in accordance with the contrast density, occasionally requiring editing by analysts. The following parameters are obtained through QCA: (i) minimal lumen diameter; (ii) reference vessel diameter; (iii) obstruction length, (iv) and percent diameter stenosis.

TIMI flow, myocardial blush, and the corrected TIMI frame count grades in the infarcted vessel were defined as previously reported.^29–31^ Each lesion with ≥50% diameter stenosis in vessels ≥1.5 mm in diameter was scored using the syntax score algorithm (https://syntaxscore.org/) fully described elsewhere^32^.

### Cardiac Magnetic Resonance Imaging

In 3.0 T clinical scanners (Siemens, Erlangen, Germany, Philips) patients remained in supine position and a phased-array receiver cardiac coil was placed on the chest. Cine images were ECG-gated and were acquired with approximately 8 sec breath holds for each slice. Cine images were acquired in the short-axis view (from the mitral-valve–insertion plane through the left ventricle until the apex) and three long-axis views. Slice thickness was 8 mm and slice spacing was 2 mm. All indexes studies were done with the injection of a commercially available gadolinium-based contrast agent (gadopentetate dimeglumine or gadoteridol) administered intravenously at a dose of 0.2 mmol per kilogram of body weight, and a dynamic, breath-hold MR first-pass perfusion examination were obtained and resulted in three short-axis slices of 8–10 mm thickness were acquired every heart beat along the long axis, with spatial resolution of 2–3 × 2–3 mm. Stress data were acquired following 6 min of dipyridamole (0.56 mg/min/kg intravenous), followed by an aminophylline injection in order to revert hyperemia. A rest injection of the same contrast agent (0.2 mmol per kilogram of body weight) was done to obtain images that would be compared with those of the stress phase. After a 5- minute delay to allow for washout of the GBCA from healthy myocardium, magnitude and phase-sensitive inversion recovery images following and inversion recovery pulse were recorded to allow the detection of late gadolinium enhancement (LGE). These were registered in the same views used for cine MRI and used to assess for myocardial viability. The inversion time suggested by protocol could be adjusted by the technologist to try to optimize image quality. Typical voxel size was 1.9 by 1.4 by 8 mm. Scanning protocol for the follow up study included segmented cine CMR (short and long axis views) for cardiac function followed by segmented LGE imaging for viability.

### Study endpoints and predictor definitions

The study endpoints were the comparison at 30 days treatment with ticagrelor or clopidogrel for the amount of myocardial infarcted mass and LVEF measured by cardiac MRI among STEMI patients under pharmaco-invasive strategy. Independent clinical, inflammatory or biochemical variables were also examined.

### Statistical analysis

All analyses were made using software R Core Team (2024) and the level of significance was stated at 5%. Data was reported as median and interquartile range or mean and standard deviation. Qualitative variables were reported as number and percentage. The Shapiro-Wilk test was used to examine normality. The Mann-Whitney test was used to compare non parametric variables for independent groups, including myocardial fibrosis (% and g), and left ventricular ejection fraction (LVEF). Categorical variables were compared by the Pearson’s Chi square test. Correlations between variables were examined by the Spearman’s Rho test. The K-means algorithm was used for cluster analysis involving LVEF and myocardial fibrosis.

## RESULTS

Figure 1 shows the study flow chart. A total of 286 patients were assessed for eligibility and from those enrolled in the trial only few patients did not complete the trial or did not perform the cMRI studies.

**Figure 1.**
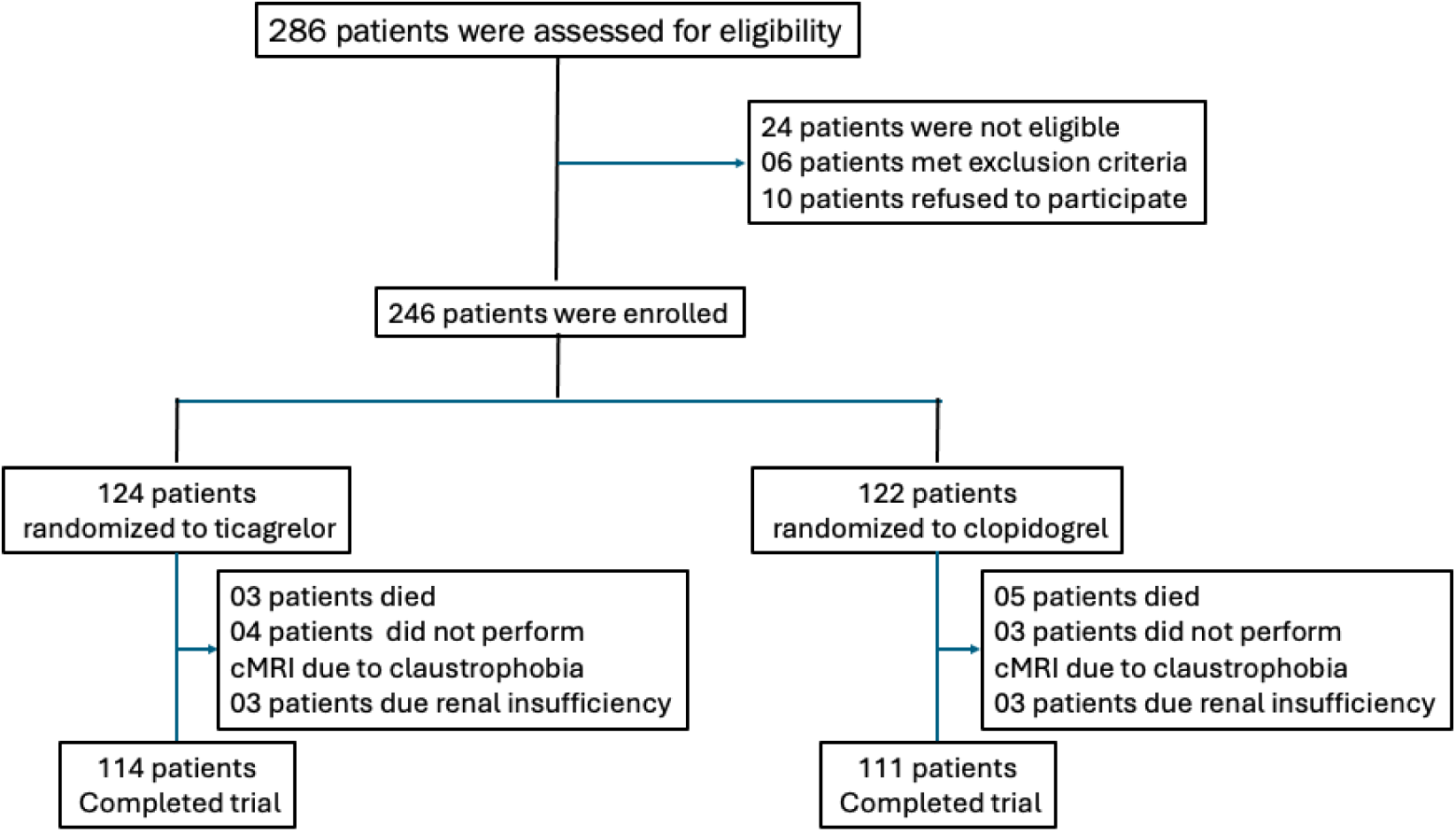
Flowchart. Of the 286 patients assessed, 40 were not enrolled in the trial due to inclusion/exclusion criteria or refusal to participate. Among those randomized to ticagrelor or clopidogrel, 21 did not complete the trial.

### Baseline Characteristics

Table 1 shows the major characteristics of the study population, composed mainly by middle aged men, a quarter of whom had diabetes and were smokers. Among laboratory parameters, higher levels of high-sensitivity troponin T (hsTNT) and high-sensitivity C-reactive protein (hsCRP) were observed among the patients randomized to the clopidogrel arm in the first day after hospitalization (Figure 2). All patients were weekly followed to receive optimal medical therapy (91% received betablockers, 97% ACEI or ARB, 100% statins monotherapy or combined with ezetimibe), and no differences were noted for clinical therapy between antiplatelet arms. Median (IQR) time for pharmacological thrombolysis was 199 (120-270) minutes and the median time from thrombolysis to percutaneous coronary intervention was 18 (9-25) hours, without differences between the arms of the study. All these interventions conform to the recommended guidelines^33^.

**Figure 2.**
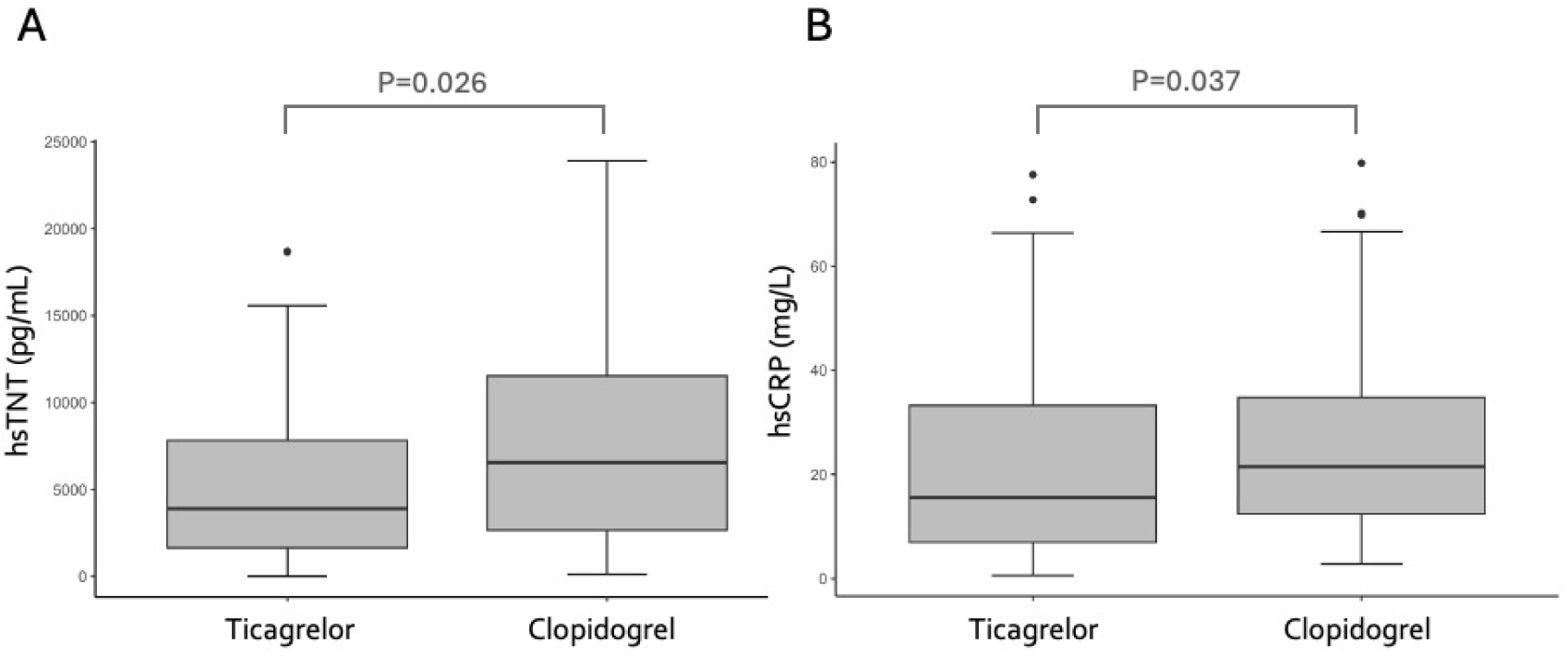
Box plots illustrating differences between the ticagrelor and clopidogrel groups on the first day of myocardial infarction. **A** – High- sensitivity troponin (hsTNT); **B** – High-sensitivity C-reactive protein (hsCRP).

**Table 1.**
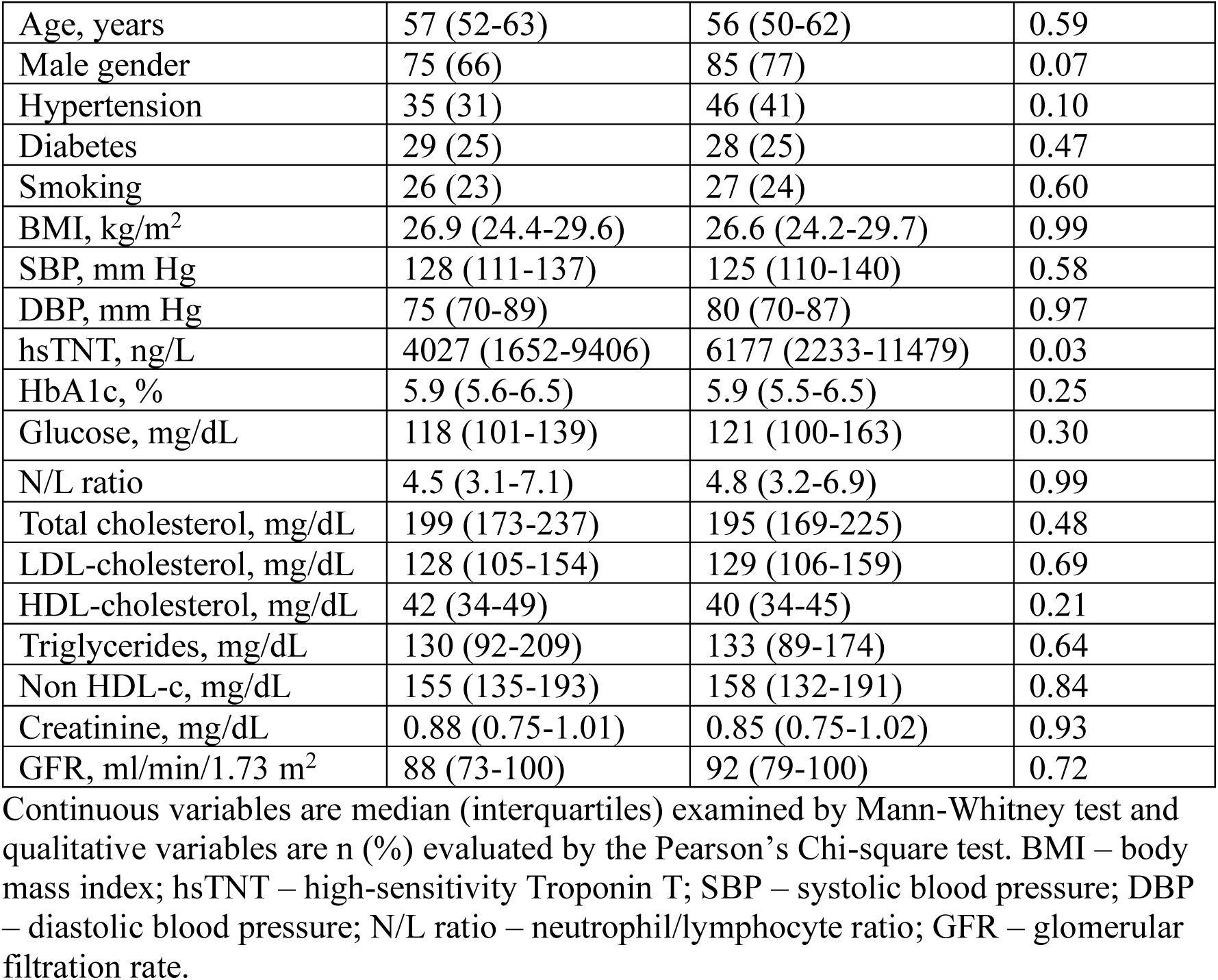
Characteristics of study population at baseline.

### Laboratory parameters

After 30 days, no differences between antiplatelet arms were observed for the standard lipid panel (total cholesterol, LDL-C, HDL-C, triglycerides, non-HDL-C), as well as for glucose, creatinine, or for inflammatory variables, including lymphocyte subtypes and interleukins (Table 2).

**Table 2.** Inflammatory markers by treatment groups at baseline and 30 days.

|  | Ticagrelor | Clopidogrel | P value |
| --- | --- | --- | --- |
| **At baseline** |  |  |  |
| N/L ratio | 4.5 (3.1-7.1) | 4.7 (3.2-7.0) | 0.99 |
| HsCRP, mg/L | 17.8 (7.0-40.3) | 23.4 (12.7-47.2) | 0.03 |
| TCD4, cells/mL | 960 (611-1294) | 869 (558-1228) | 0.51 |
| TCD8, cells/mL | 352 (243-442) | 315 (212-533) | 0.83 |
| B1, cells/mL | 4.7 (2.1-10.2) | 4.1 (2.5-7.8) | 0.67 |
| B2 mem, cells/mL | 57 (28-122) | 48 (24-110) | 0.74 |
| B2 naïve, cells/mL | 65 (14-118) | 48 (17-102) | 0.99 |
| B2 class, cells/mL | 138 (87-260) | 124 (71-221) | 0.55 |
| IL-1b, pg/mL | 0.06 (0.00-0.12) | 0.05 (0.03-0.12) | 0.54 |
| IL-4, pg/mL | 3.7 (0.0-8.5) | 1.7 (0.0-8.1) | 0.27 |
| IL-6, pg/mL | 6.2 (0.0-20.0) | 7.1 (0.0-29.0) | 0.48 |
| IL-10, pg/mL | 4.5 (0.0-11.8) | 3.3 (0.2-10.1) | 0.76 |
| IL-18, pg/mL | 1145 (696-1600) | 1123 (872-1653) | 0.55 |
| **At 30 days** |  |  |  |
| N/L ratio | 2.8 (2.4-3.6) | 2.9 (2.2-3.7) | 0.73 |
| hsCRP, mg/L | 2.1 (1.1-4.3) | 2.3 (0.9-6.4) | 0.92 |
| TCD4, cells/mL | 841 (685-1095) | 886 (656-1203) | 0.83 |
| TCD8, cells/mL | 329 (212-506) | 282 (198-498) | 0.82 |
| B1, cells/mL | 4.1 (2.5-9.1) | 3.0 (1.9-7.1) | 0.09 |
| B2 mem, cells/mL | 48 (26-96) | 38 (20-81) | 0.28 |
| B2 naïve, cells/mL | 41 (14-79) | 41 (11-87) | 0.94 |
| B2 class, cells/mL | 108 (67-172) | 100 (52-149) | 0.49 |
| IL-1b, pg/mL | 0.02 (0.00-0.09) | 0.02 (0.00-0.11) | 0.75 |
| IL-4, pg/mL | 6.7 (3.5-10.7) | 6.0 (2.1-9.8) | 0.53 |
| IL-6, pg/mL | 11.2 (8.9-15.6) | 12.1 (8.8-14.9) | 0.96 |
| IL-10, pg/mL | 17.2 (7.3-26.5) | 16.9 (5.3-30.9) | 0.88 |
| IL-18, pg/mL | 351 (207-482) | 346 (216-463) | 0.80 |

Values are median (interquartiles). Variables were tested by the Mann-Whitney U test. N/L – neutrophil/lymphocyte; hsCRP – high-sensitivity C-reactive protein; B2 mem – B2 memory; B2 class – B2 classic (B naïve plus B memory cells).

### Angiographic findings and PCI details

Coronary angiography and percutaneous coronary intervention were performed at the first 24 hours of STEMI. All patients received the randomized antiplatelet therapy before the hemodynamic procedures.

The culprit coronary arteries were predominantly left anterior descending artery (LAD) or right coronary artery (RCA) and less frequently left circumflex (LCx). There were no differences between arms according to the type of culprit artery (p=0.77, Chi-square test). The SYNTAX score was similar between arms [ticagrelor 6.5 (4.0-9.5), clopidogrel 5.5 (4.0-10.0), p=0.62, Mann-Whitney test]. The Gensini score was also similar between arms [ticagrelor 10 (4-21); clopidogrel 8 (4-18), p=0.47, Mann-Whitney test]. There were no differences for the TIMI flow grade (0, 1, 2, or 3) pre (p=0.78) or post PCI (p=0.58; Chi-square test), being in both arms predominantly TIMI grade 3.

The majority of patients achieved myocardial blush grade 3 in both arms (p=0.90, Chi-square test). In addition, few patients had RENTROP score 1 or 2, with the vast majority not showing collateral vessels, with no differences between arms (p=0.40, Chi-square test).

### Cardiac MRI outcomes

Smaller amount of infarcted mass was observed in the ticagrelor arm compared with clopidogrel arm in grams (p=0.016, Mann-Whitney test) or in the percentage of left ventricular mass (p=0.012, Mann-Whitney test). The left ventricular mass did not differ between groups (p=0.57, Mann-Whitney test) as well as for the left ventricular ejection fraction (LVEF) (p=0.06, Mann-Whitney test).

Exploratory correlations with myocardial fibrosis at 1^st^ day and 30^th^ day are shown by network graph in the ticagrelor and clopidogrel arms (Figure 3). Network graph shows the correlations with LVEF by treatments (Figure 4).

**Figure 3.**
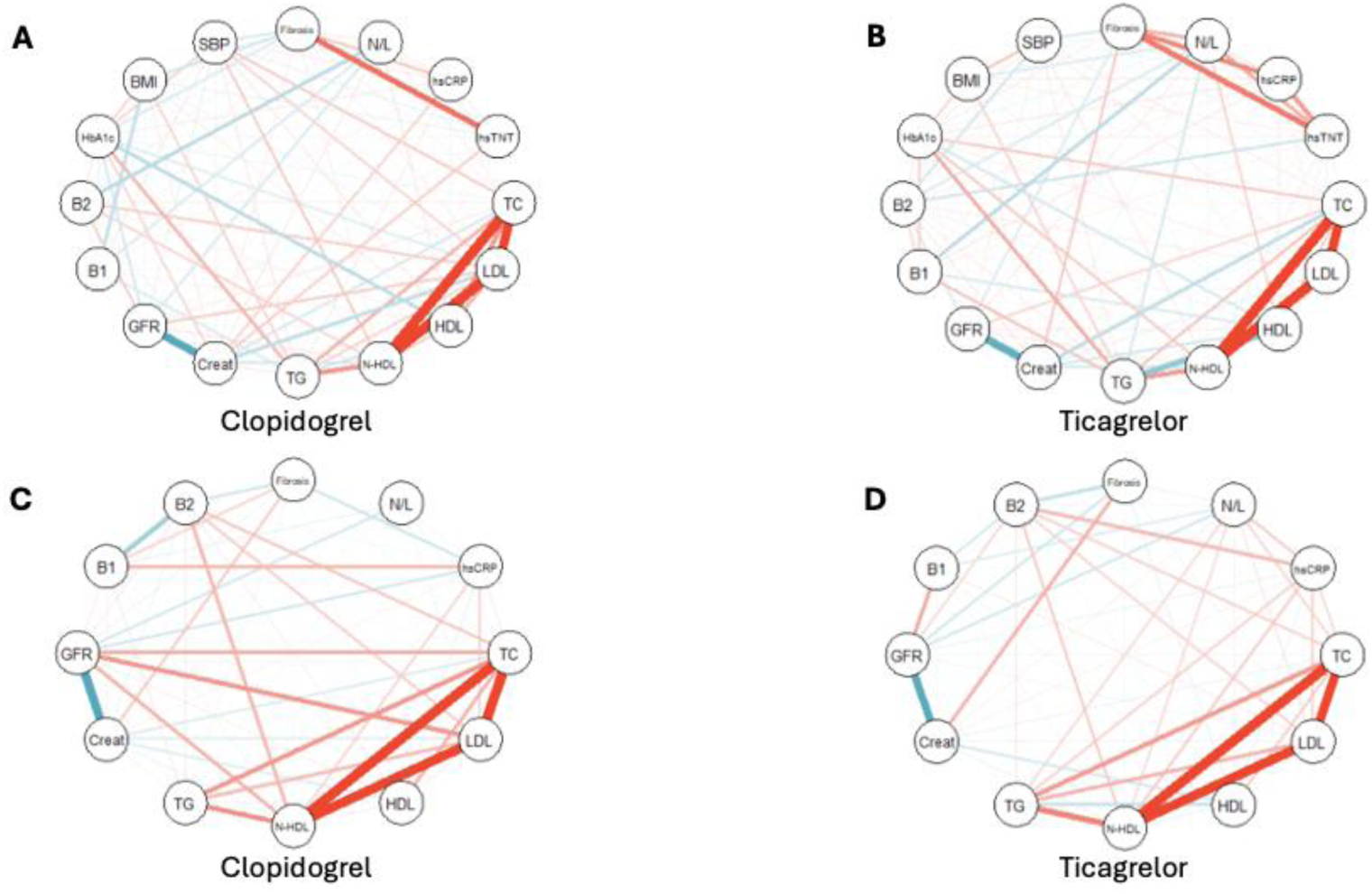
Network graph showing variable correlations on the **1st day** (A, B) and the **30th day** (C, D). In the ticagrelor arm, the most significant correlations with myocardial fibrosis on day 1 were high-sensitivity troponin T (hsTNT) and high-sensitivity C-reactive protein (hsCRP), whereas in the clopidogrel arm, hsTNT was the primary variable. By day 30, hsCRP, B2 cells, and creatinine were the main variables correlated with fibrosis in the clopidogrel arm, while creatinine and B2 cells were predominant in the ticagrelor arm. **Blue lines** indicate negative correlations; **red lines** indicate positive correlations.

**Figure 4.**
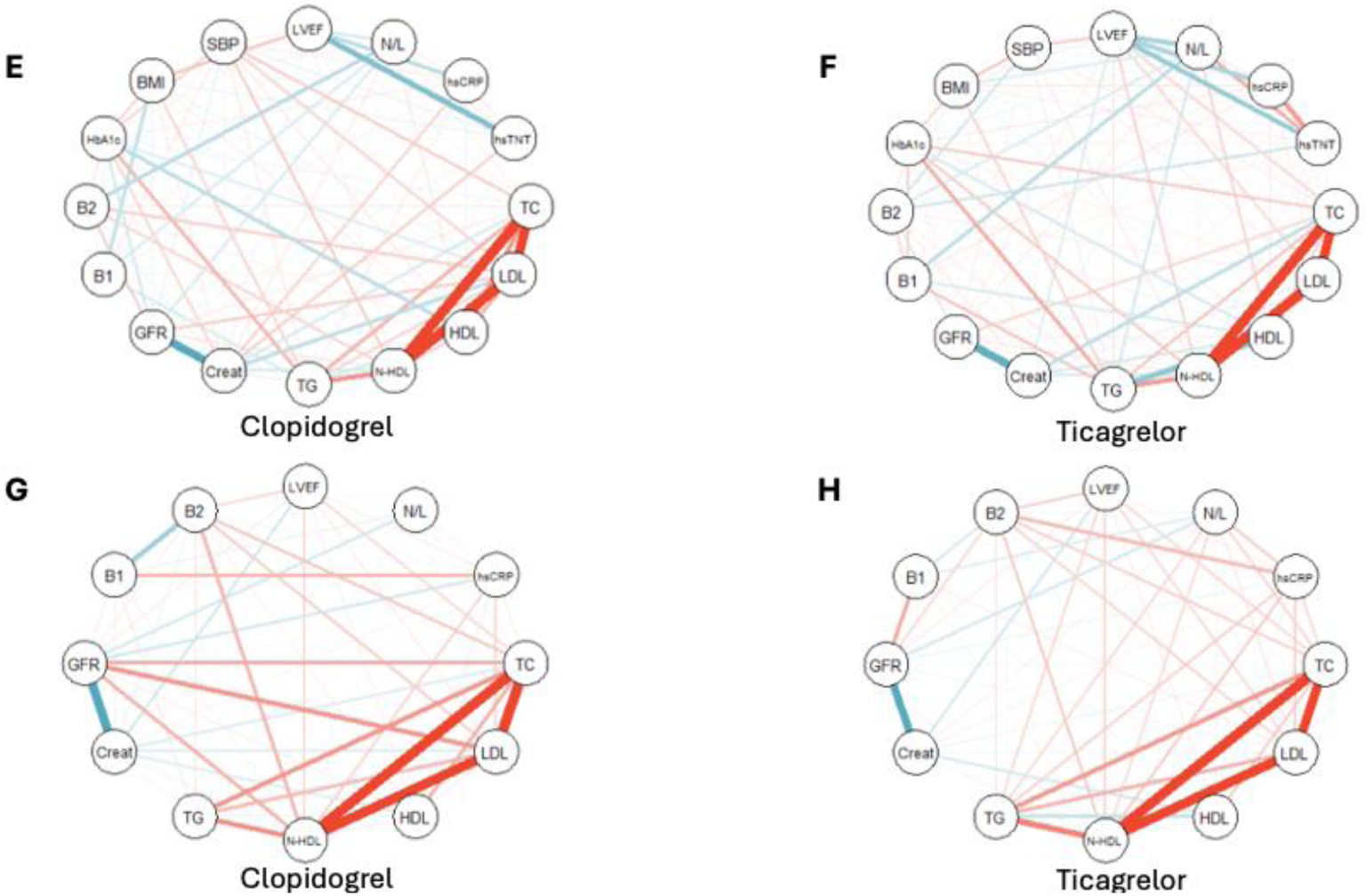
Network graph depicting the most important correlations with left ventricular ejection fraction (LVEF). On the **1st day** (E, F), in the clopidogrel arm, key correlations included high-sensitivity troponin T (hsTNT), high-sensitivity C-reactive protein (hsCRP), neutrophil-to-lymphocyte ratio (N/L), and systolic blood pressure (SBP), while in the ticagrelor arm, the same variables, along with lipid parameters, were correlated with LVEF. By the **30th day** (G, H), in the clopidogrel arm, creatinine, B2 cells, and lipid parameters were the main correlations with LVEF, whereas in the ticagrelor arm, B2 cells, creatinine, and lipid parameters were also predominant. **Blue lines** indicate negative correlations; **red lines** indicate positive correlations.

The K-means algorithm shows data obtained for fibrosis (%) and LVEF in clusters for ticagrelor and clopidogrel (Figure 5). The accuracy obtained was 57%, showing, in the ticagrelor arm, more patients displaying lower infarcted mass and higher LVEF.

**Figure 5.**
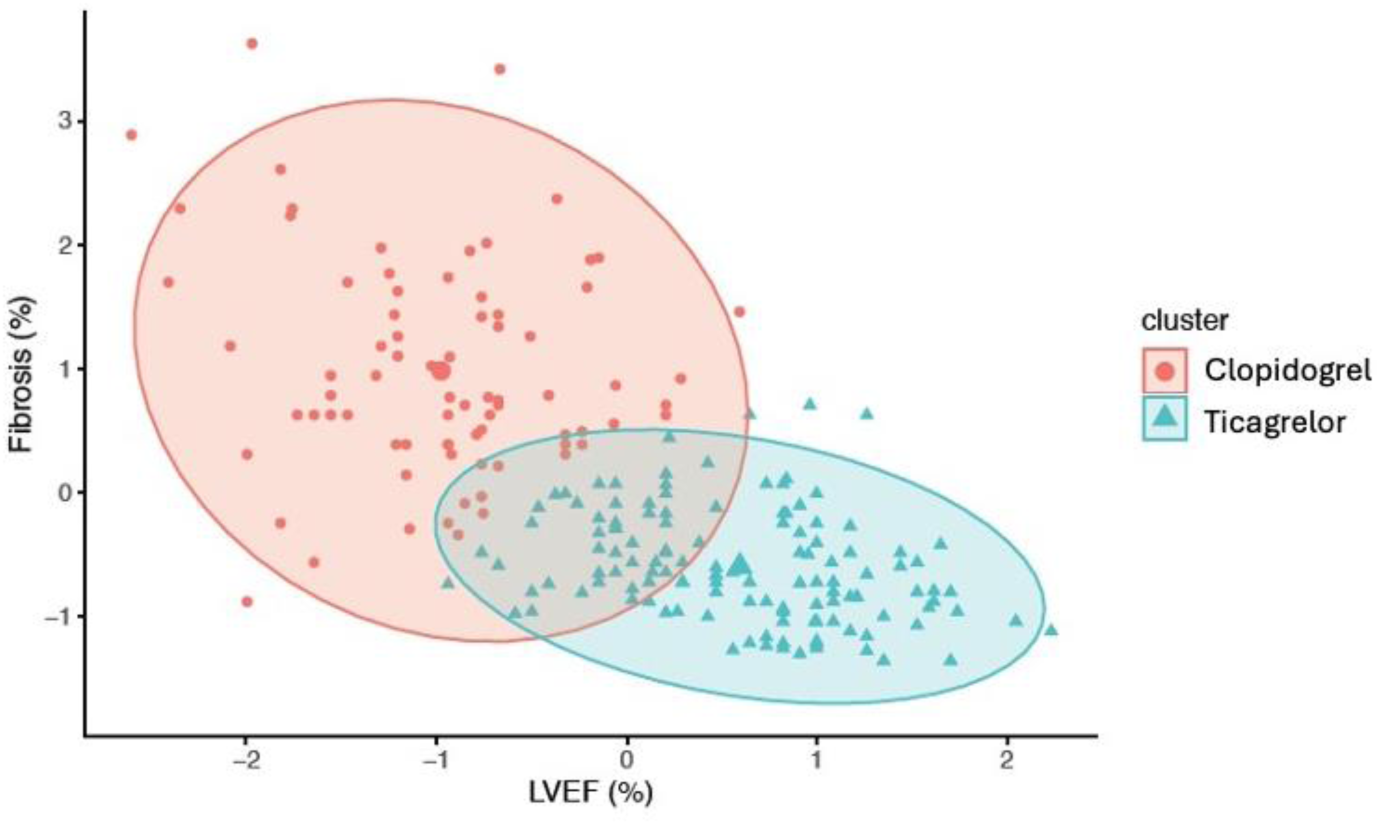
Clusters of fibrosis and LVEF in clopidogrel and ticagrelor arms. K-means analysis demonstrates moderate accuracy (57.3%). The graph shows that most patients in the ticagrelor arm are clustered with better LVEF and smaller infarct size.

### Clinical Outcomes

The study was not powered to evaluate clinical events. There were four patients hospitalized with the need of urgent PCI, one patient with recurrent myocardial infarction and one hospitalized due to heart failure. Six patients were submitted to elective CABG and 22 to elective PCI. During the first month post STEMI, eight patients died (three in the ticagrelor arm and five in the clopidogrel arm). No differences between arms were observed for these clinical events. The trial was stopped on March 2020 after reaching the estimated sample size for the primary objective of the BATTLE- AMI study^24^.

## DISCUSSION

The main contribution of this study was to show that antiplatelet therapy with ticagrelor in a randomized clinical trial, compared with clopidogrel, was associated with smaller infarcted mass by cMRI 30 days after STEMI. The study also revealed that patients treated with ticagrelor had more homogeneous response towards smaller infarcted mass and better left ventricular function.

All patients were randomized to receive antiplatelet therapy prior to invasive procedures, within an appropriate window for thrombolytic therapy (less than six hours from symptom onset). Additionally, coronary angiography followed by percutaneous coronary interventions was performed within the first 24 hours of STEMI. A third key feature of the study was the implementation of rigorous, weekly follow-ups after hospital discharge to optimize clinical therapies.

The study was designed with the hypothesis that ticagrelor, by reducing adenosine uptake into cells, could enhance microcirculation and thereby promote the recovery of ischemic cardiomyocytes^24^. Indeed, the improvement of coronary microcirculation with ticagrelor has been previously demonstrated in patients undergoing elective percutaneous coronary intervention, using continuous thermodilution to analyze coronary blood flow and microvascular resistance at baseline and after antiplatelet therapy^34^. The lower hsTNT levels observed on the first day of STEMI in the ticagrelor group suggest an early benefit on microcirculation, as both study arms had a similar distribution of culprit arteries, SYNTAX, and Gensini scores after randomization. Furthermore, post-PCI assessments, including TIMI flow, myocardial blush, and Rentrop scores, were also comparable between the two groups. Collectively, this early improvement in microcirculation may have contributed, at least in part, to the reduction in myocardial injury.

Reduction in inflammation has been proposed as a mechanism for improving cardiovascular risk^14,15^. Additionally, inflammatory markers have been associated with infarct size and ventricular function following STEMI^25,26^. In our study, patients randomized to the clopidogrel arm exhibited higher hsCRP levels measured on the first day of STEMI. A previous study investigated inflammatory markers (hsCRP, interleukin-6, and endothelial cell-specific molecule 1) in STEMI patients randomized to treatment with clopidogrel or ticagrelor. These markers were assessed at admission, 24 hours, day 4, and day 7 post-STEMI. The authors reported that, compared to clopidogrel, ticagrelor appeared to reduce these inflammatory markers more rapidly^35^. Thus, the differences observed in hsCRP levels between the ticagrelor and clopidogrel groups in our study may be attributed to a faster anti-inflammatory response with ticagrelor following STEMI^35^. A lower inflammatory state could also contribute to a smaller infarct size observed in the ticagrelor arm.

More effective lipid-lowering therapy has consistently been associated with clinical benefits and stabilization of atherosclerotic plaques^36^. In our study, all patients received high-intensity lipid-lowering therapy, with comparable standard lipid panel between arms at baseline and after 30 days. However, a prior study explored the effects of antiplatelet therapy on LDL particle quality using Gaussian laser beam (Z-scan), small-angle X-ray scattering (SAXS), dynamic light scattering (DLS), ultraviolet-visible spectroscopy, and polyacrylamide gel electrophoresis. These methods assessed oxidative status, structural changes, particle size, and LDL subfractions, revealing better LDL quality in samples from patients treated with ticagrelor compared to clopidogrel^37^. The affinity of LDL receptors is influenced by LDL particle size and oxidation status, and the improved LDL quality following ticagrelor therapy may have long-term relevance. However, network analysis indicated weak short-term correlations between standard lipid parameters and both infarct size and left ventricular ejection fraction (LVEF).

### Study limitations and strengths

The BATTLE-AMI trial included only STEMI patients undergoing pharmaco- invasive therapy, therefore, caution is needed when extrapolating these findings to patients treated with primary angioplasty. Despite observing differences in infarct size, there was only a trend toward improved left ventricular ejection fraction in the ticagrelor arm, likely due to the optimization of clinical therapies (including beta-blockers, renin-angiotensin system blockers, and lipid-lowering therapies), as well as the timely thrombolysis and coronary interventions.

## CONCLUSIONS

Patients with STEMI who received optimal medical therapy under a pharmaco- invasive strategy, with invasive procedures performed within an appropriate time window, showed that those randomized to ticagrelor had a smaller infarct size as measured by cMRI compared to clopidogrel, despite similar pre- and post-reperfusion angiographic characteristics of coronary disease.

## Data Availability

Data referred to in the manuscript can be available upon authorization of the authors.

## Author Contributions

FAF and MCI are the principal investigators of this trial and drafted the manuscript. FAF obtained the financial support, FAF and MCI participated in the study design, and carried out patient recruitment. IP and GZ participated in the cMRI analysis. AFN participated in the lipid analysis. CF performed flow-cytometry studies. HB and AB carried out patient recruitment and follow-up. HF participated in the analysis of inflammatory parameters. AC, MB and IB participated in the coronary angiographic analysis. All authors read and approved the manuscript.

## Acknowledgments

We thank Mayara M Garcia (BioStats, Brazil) for the statistical analysis of the study.

## References

1. Wallentin L, Becker RC, Budaj A, Cannon CP, Emanuelsson H, Held C, Horrow J, Husted S, James S, Katus H, Mahaffey KW, Scirica BM, Skene A, Steg PG, Storey RF, Harrington RA; PLATO Investigators, Freij A, Thorsén M. Ticagrelor versus clopidogrel in patients with acute coronary syndromes. N Engl J Med. 2009;361(11):1045–57. doi: 10.1056/NEJMoa0904327.

2. Jeong YJ, Park K, Kim YD. Comparison between ticagrelor and clopidogrel on myocardial blood flow in patients with acute coronary syndrome, using 13 N-ammonia positron emission tomography. Am Heart J. 2020;222:121–130. doi: 10.1016/j.ahj.2020.01.013.

3. Park K, Cho YR, Park JS, Park TH, Kim MH, Kim YD. Comparison of the Effects of ticagrelor and clopidogrel on microvascular dysfunction in patients with acute coronary syndrome using invasive physiologic indices. Circ Cardiovasc Interv. 2019;12(10):e008105. doi: 10.1161/CIRCINTERVENTIONS.119.008105.

4. Gimbel M, Qaderdan K, Willemsen L, Hermanides R, Bergmeijer T, de Vrey E, Heestermans T, Tjon Joe Gin M, Waalewijn R, Hofma S, den Hartog F, Jukema W, von Birgelen C, Voskuil M, Kelder J, Deneer V, Ten Berg J. Clopidogrel versus ticagrelor or prasugrel in patients aged 70 years or older with non-ST-elevation acute coronary syndrome (POPular AGE): the randomised, open-label, non-inferiority trial. Lancet. 2020;395(10233):1374-1381. doi: 10.1016/S0140-6736(20)30325-1.

5. Zhao X, Zhang J, Guo J, Wang J, Pan Y, Zhao X, Sang W, Yang K, Xu F, Xu F, Chen Y. Comparison of Safety and Efficacy Between Clopidogrel and Ticagrelor in Elderly Patients With Acute Coronary Syndrome: A Systematic Review and Meta-Analysis. Front Pharmacol. 2021;12:743259. doi: 10.3389/fphar.2021.743259.

6. Verdoia M, Savonitto S, Dudek D, Kedhi E, De Luca G. Ticagrelor as compared to conventional antiplatelet agents in coronary artery disease: A comprehensive meta-analysis of 15 randomized trials. Vascul Pharmacol. 2021;137:106828. doi: 10.1016/j.vph.2020.106828.

7. Valgimigli M, Gragnano F, Branca M, Franzone A, da Costa BR, Baber U, Kimura T, Jang Y, Hahn JY, Zhao Q, Windecker S, Gibson CM, Watanabe H, Kim BK, Song YB, Zhu Y, Vranckx P, Mehta S, Ando K, Hong SJ, Gwon HC, Serruys PW, Dangas GD, McFadden EP, Angiolillo DJ, Heg D, Calabrò P, Jüni P, Mehran R; Single Versus Dual Antiplatelet Therapy (Sidney-3) Collaboration. Ticagrelor or Clopidogrel Monotherapy vs Dual Antiplatelet Therapy After Percutaneous Coronary Intervention: A Systematic Review and Patient-Level Meta-Analysis. JAMA Cardiol. 2024 May 1;9(5):437-448. doi: 10.1001/jamacardio.2024.0133.

8. Ge Z, Kan J, Gao X, Raza A, Zhang JJ, Mohydin BS, Gao F, Shao Y, Wang Y, Zeng H, Li F, Khan S, Mengal N, Cong H, Wang M, Chen L, Wei Y, Chen F, Stone GW, Chen SL, ULTIMATE-DAPT investigators. Lancet. 2024 May 11;403(10439):1866-1878. doi: 10.1016/S0140-6736(24)00473-2.

9. Berwanger O, Nicolau JC, Carvalho AC, Jiang L, Goodman SG, Nicholls SJ, Parkhomenko A, Averkov O, Tajer C, Malaga G, Saraiva JFK, Fonseca FA, De Luca FA, Guimaraes HP, de Barros E, Silva PGM, Damiani LP, Paisani DM, Lasagno CMR, Candido CT, Valeis N, Moia DDF, Piegas LS, Granger CB, White HD, Lopes RD; TREAT Study Group. Ticagrelor vs Clopidogrel After Fibrinolytic Therapy in Patients With ST-Elevation Myocardial Infarction: A Randomized Clinical Trial. JAMA Cardiol. 2018;3(5):391–399. doi: 10.1001/jamacardio.2018.0612.

10. Berwanger O, Lopes RD, Moia DDF, Fonseca FA, Jiang L, Goodman SG, Nicholls SJ, Parkhomenko A, Averkov O, Tajer C, Malaga G, Saraiva JFK, Guimaraes HP, de Barros E Silva PGM, Damiani LP, Santos RHN, Paisani DM, Miranda TA, Valeis N, Piegas LS, Granger CB, White HD, Nicolau JC. Ticagrelor Versus Clopidogrel in Patients With STEMI Treated With Fibrinolysis: TREAT Trial. J Am Coll Cardiol. 2019;73(22):2819–2828. doi: 10.1016/j.jacc.2019.03.011.

11. Cannon CP, Blazing MA, Giugliano RP, McCagg A, White JA, Theroux P, Darius H, Lewis BS, Ophuis TO, Jukema JW, De Ferrari GM, Ruzyllo W, De Lucca P, Im K, Bohula EA, Reist C, Wiviott SD, Tershakovec AM, Musliner TA, Braunwald E, Califf RM; IMPROVE-IT Investigators. Ezetimibe Added to Statin Therapy after Acute Coronary Syndromes. N Engl J Med. 2015;372(25):2387–97. doi: 10.1056/NEJMoa1410489.

12. Schwartz GG, Gabriel Steg P, Bhatt DL, Bittner VA, Diaz R, Goodman SG, Jukema JW, Kim YU, Li QH, Manvelian G, Pordy R, Sourdille T, White HD, Szarek M; ODYSSEY OUTCOMES Committees and Investigators. Clinical Efficacy and Safety of Alirocumab After Acute Coronary Syndrome According to Achieved Level of Low-Density Lipoprotein Cholesterol: A Propensity Score-Matched Analysis of the ODYSSEY OUTCOMES Trial. Circulation. 2021;143(11):1109–1122. doi: 10.1161/CIRCULATIONAHA.120.049447.

13. Ridker PM, Danielson E, Fonseca FA, Genest J, Gotto AM Jr, Kastelein JJ, Koenig W, Libby P, Lorenzatti AJ, MacFadyen JG, Nordestgaard BG, Shepherd J, Willerson JT, Glynn RJ; JUPITER Study Group. Rosuvastatin to prevent vascular events in men and women with elevated C-reactive protein. N Engl J Med. 2008;359(21):2195–207. doi: 10.1056/NEJMoa0807646.

14. Ridker PM, Everett BM, Thuren T, MacFadyen JG, Chang WH, Ballantyne C, Fonseca F, Nicolau J, Koenig W, Anker SD, Kastelein JJP, Cornel JH, Pais P, Pella D, Genest J, Cifkova R, Lorenzatti A, Forster T, Kobalava Z, Vida-Simiti L, Flather M, Shimokawa H, Ogawa H, Dellborg M, Rossi PRF, Troquay RPT, Libby P, Glynn RJ; CANTOS Trial Group. Antiinflammatory Therapy with Canakinumab for Atherosclerotic Disease. N Engl J Med. 2017;377(12):1119–1131. doi: 10.1056/NEJMoa1707914.

15. Tardif JC, Kouz S, Waters DD, Bertrand OF, Diaz R, Maggioni AP, Pinto FJ, Ibrahim R, Gamra H, Kiwan GS, Berry C, López-Sendón J, Ostadal P, Koenig W, Angoulvant D, Grégoire JC, Lavoie MA, Dubé MP, Rhainds D, Provencher M, Blondeau L, Orfanos A, L’Allier PL, Guertin MC, Roubille F. Efficacy and Safety of Low-Dose Colchicine after Myocardial Infarction. N Engl J Med. 2019;381(26):2497–2505. doi: 10.1056/NEJMoa1912388.

16. Mega JL, Braunwald E, Wiviott SD, Bassand JP, Bhatt DL, Bode C, Burton P, Cohen M, Cook-Bruns N, Fox KA, Goto S, Murphy SA, Plotnikov AN, Schneider D, Sun X, Verheugt FW, Gibson CM; ATLAS ACS 2–TIMI 51 Investigators. Rivaroxaban in patients with a recent acute coronary syndrome. N Engl J Med. 2012 Jan 5;366(1):9–19. doi: 10.1056/NEJMoa1112277.

17. Bonaca MP, Bhatt DL, Cohen M, Steg PG, Storey RF, Jensen EC, Magnani G, Bansilal S, Fish MP, Im K, Bengtsson O, Oude Ophuis T, Budaj A, Theroux P, Ruda M, Hamm C, Goto S, Spinar J, Nicolau JC, Kiss RG, Murphy SA, Wiviott SD, Held P, Braunwald E, Sabatine MS; PEGASUS-TIMI 54 Steering Committee and Investigators. Long-term use of ticagrelor in patients with prior myocardial infarction. N Engl J Med. 2015;372(19):1791–800. doi: 10.1056/NEJMoa1500857.

18. Cesaro A, Gragnano F, Calabrò P, Moscarella E, Santelli F, Fimiani F, Patti G, Cavallari I, Antonucci E, Cirillo P, Pignatelli P, Palareti G, Pelliccia F, Bossone E, Pengo V, Gresele P, Marcucci R; START-ANTIPLATELET collaborators. Prevalence and clinical implications of eligibility criteria for prolonged dual antithrombotic therapy in patients with PEGASUS and COMPASS phenotypes: Insights from the START-ANTIPLATELET registry. Int J Cardiol. 2021;345:7–13. doi: 10.1016/j.ijcard.2021.10.138.

19. Klingenberg R, Aghlmandi S, Räber L, Gencer B, Nanchen D, Heg D, Carballo S, Rodondi N, Mach F, Windecker S, Jüni P, von Eckardstein A, Matter CM, Lüscher TF. Improved risk stratification of patients with acute coronary syndromes using a combination of hsTnT, NT-proBNP and hsCRP with the GRACE score. Eur Heart J Acute Cardiovasc Care. 2018;7(2):129–138. doi: 10.1177/2048872616684678.

20. Stone GW, Selker HP, Thiele H, Patel MR, Udelson JE, Ohman EM, Maehara A, Eitel I, Granger CB, Jenkins PL, Nichols M, Ben-Yehuda O. Relationship Between Infarct Size and Outcomes Following Primary PCI: Patient-Level Analysis From 10 Randomized Trials. J Am Coll Cardiol. 2016;67(14):1674–83. doi: 10.1016/j.jacc.2016.01.069.

21. Sinnaeve PR, Armstrong PW, Gershlick AH, Goldstein P, Wilcox R, Lambert Y, Danays T, Soulat L, Halvorsen S, Ortiz FR, Vandenberghe K, Regelin A, Bluhmiki E, Bogaerts K, Van de Werf F; STREAM investigators. ST-segment-elevation myocardial infarction patients randomized to a pharmaco-invasive strategy or primary percutaneous coronary intervention: Strategic Reperfusion Early After Myocardial Infarction (STREAM) 1-year mortality follow-up. Circulation. 2014; 30(14):1139-45. doi: 10.1161/CIRCULATIONAHA.114.009570.

22. Siddiqi TJ, Usman MS, Khan MS, Sreenivasan J, Kassas I, Riaz H, et al. Meta-Analysis Comparing Primary Percutaneous Coronary Intervention Versus Pharmacoinvasive Therapy in Transfer Patients with ST-Elevation Myocardial Infarction. Am J Cardiol. 2018;122(4):542–7. doi: 10.1016/j.amjcard.2018.04.057.

23. Shavadia J, Zheng Y, Dianati Maleki N, Huber K, Halvorsen S, Goldstein P, et al. Infarct Size, Shock, and Heart Failure: Does Reperfusion Strategy Matter in Early Presenting Patients With ST-Segment Elevation Myocardial Infarction? J Am Heart Assoc. 2015;4(8):e002049. doi: 10.1161/JAHA.115.002049.

24. Fonseca FAH, Izar MC, Maugeri IML, Berwanger O, Damiani LP, Pinto IM, Szarf G, França CN, Bianco HT, Moreira FT, Caixeta A, Alves CMR, Soriano Lopes A, Klassen A, Tavares MFM, Fonseca HA, Carvalho ACC; BATTLE-AMI Investigators. Effects of four antiplatelet/statin combined strategies on immune and inflammatory responses in patients with acute myocardial infarction undergoing pharmacoinvasive strategy: Design and rationale of the B and T Types of Lymphocytes Evaluation in Acute Myocardial Infarction (BATTLE-AMI) study: study protocol for a randomized controlled trial. Trials. 2017;18(1):601. doi: 10.1186/s13063-017-2361-1.

25. Casarotti ACA, Teixeira D, Longo-Maugeri IM, Ishimura ME, Coste MER, Bianco HT, Moreira FT, Bacchin AF, Izar MC, Gonçalves I, Caixeta A, Szarf G, Pinto IM, Fonseca FA. Role of B lymphocytes in the infarcted mass in patients with acute myocardial infarction. Biosci Rep. 2021;41(2):BSR20203413. doi: 10.1042/BSR20203413.

26. Coste MER, França CN, Izar MC, Teixeira D, Ishimura ME, Longo-Maugeri I, Bacchin AS, Bianco HT, Moreira FT, Pinto IM, Szarf G, Caixeta AM, Berwanger O, Gonçalves I Jr, Fonseca FAH. Early Changes in Circulating Interleukins and Residual Inflammatory Risk After Acute Myocardial Infarction. Arq Bras Cardiol. 2020;115(6):1104–11. doi: 10.36660/abc.20190567.

27. Qualitative and Quantitative Coronary Angiography. In Eric Topol: Textbook of Interventional Cardiology, 64, 999–1017.e5. 2020 by Elsevier, Inc.

28. Suzuki N, Asano T, Nakazawa G, Aoki J, Tanabe K, Hibi K, Ikari Y, Kozuma K. Clinical expert consensus document on quantitative coronary angiography from the Japanese Association of Cardiovascular Intervention and Therapeutics. Cardiovasc Interv Ther. 2020;35(2):105–116. doi: 10.1007/s12928-020-00653-7. Epub 2020 Mar 3. PMID: 32125622; PMCID: PMC7105443.

29. TIMI Study Group. The Thrombolysis in Myocardial Infarction (TIMI) trial. Phase I findings. N Engl J Med. 1985;312(14):932–6. doi: 10.1056/NEJM198504043121437. PMID: 4038784.

30. Gibson CM, Cannon CP, Daley WL, Dodge JT Jr, Alexander B Jr, Marble SJ, McCabe CH, Raymond L, Fortin T, Poole WK, Braunwald E. TIMI frame count: a quantitative method of assessing coronary artery flow. Circulation. 1996;93(5):879–88. doi: 10.1161/01.cir.93.5.879. PMID: 8598078.

31. Van ’t Hof AW, Liem A, Suryapranata H, Hoorntje JC, de Boer MJ, Zijlstra F. Angiographic assessment of myocardial reperfusion in patients treated with primary angioplasty for acute myocardial infarction: myocardial blush grade. Zwolle Myocardial Infarction Study Group. Circulation. 1998;97(23):2302-6. doi: 10.1161/01.cir.97.23.2302. PMID: 9639373.

32. Sianos G, Morel MA, Kappetein AP, Morice MC, Colombo A, Dawkins K, van den Brand M, Van Dyck N, Russell ME, Mohr FW, Serruys PW. The SYNTAX Score: an angiographic tool grading the complexity of coronary artery disease. EuroIntervention. 2005;1(2):219–27. PMID: 19758907.

33. Levine GN, Bates ER, Bittl JA, Brindis RG, Fihn SD, Fleisher LA, Granger CB, Lange RA, Mack MJ, Mauri L, Mehran R, Mukherjee D, Newby LK, O’Gara PT, Sabatine MS, Smith PK, Smith SC Jr. 2016 ACC/AHA Guideline Focused Update on Duration of Dual Antiplatelet Therapy in Patients With Coronary Artery Disease: A Report of the American College of Cardiology/American Heart Association Task Force on Clinical Practice Guidelines: An Update of the 2011 ACCF/AHA/SCAI Guideline for Percutaneous Coronary Intervention, 2011 ACCF/AHA Guideline for Coronary Artery Bypass Graft Surgery, 2012 ACC/AHA/ACP/AATS/PCNA/SCAI/STS Guideline for the Diagnosis and Management of Patients With Stable Ischemic Heart Disease, 2013 ACCF/AHA Guideline for the Management of ST-Elevation Myocardial Infarction, 2014 AHA/ACC Guideline for the Management of Patients With Non-ST-Elevation Acute Coronary Syndromes, and 2014 ACC/AHA Guideline on Perioperative Cardiovascular Evaluation and Management of Patients Undergoing Noncardiac Surgery. Circulation. 2016 2016;134(10):e123-55.doi: 10.1161/CIR.0000000000000404.

34. Mangiacapra F, Colaiori I, Di Gioia G, Pellicano M, Heyse A, Paolucci L, Peace A, Bartunek J, de Bruyne B, Barbato E. Effects of ticagrelor and prasugrel on coronary microcirculation in elective percutaneous coronary intervention. Heart. 2023;110(2):115–21. doi: 10.1136/heartjnl-2022-321868.

35. Gao CZ, Ma QQ, Wu J, Liu R, Wang F, Bai J, Yang XJ, Fu Q, Wei P. Comparison of the Effects of Ticagrelor and Clopidogrel on Inflammatory Factors, Vascular Endothelium Functions and Short-Term Prognosis in Patients with Acute ST-Segment Elevation Myocardial Infarction Undergoing Emergency Percutaneous Coronary Intervention: a Pilot Study. Cell Physiol Biochem. 2018;48(1):385–96. doi: 10.1159/000491768.

36. Räber L, Ueki Y, Otsuka T, Losdat S, Häner JD, Lonborg J, Fahrni G, Iglesias JF, van Geuns RJ, Ondracek AS, Radu Juul Jensen MD, Zanchin C, Stortecky S, Spirk D, Siontis GCM, Saleh L, Matter CM, Daemen J, Mach F, Heg D, Windecker S, Engstrøm T, Lang IM, Koskinas KC; PACMAN-AMI collaborators. Effect of Alirocumab Added to High-Intensity Statin Therapy on Coronary Atherosclerosis in Patients With Acute Myocardial Infarction: The PACMAN-AMI Randomized Clinical Trial. JAMA. 2022;327(18):1771–81. doi: 10.1001/jama.2022.5218.

37. Lotfollahi Z, Mello APQ, Fonseca FAH, Machado LO, Mathias AF, Izar MC, Damasceno NRT, Oliveira CLP, Neto AMF. Changes in lipoproteins associated with lipid-lowering and antiplatelet strategies in patients with acute myocardial infarction. PLos One. 2022;17(8):e0273292. doi: 10.1371/journal.pone.0273292.

